# Exploring the experiences of loneliness in adults with mental health problems: a participatory qualitative interview study

**DOI:** 10.1101/2022.03.02.22271346

**Authors:** Mary Birken, Beverley Chipp, Prisha Shah, Rachel Rowan Olive, Patrick Nyikavaranda, Jackie Hardy, Anjie Chhapia, Nick Barber, Stephen Lee, Eiluned Pearce, Brynmor Lloyd-Evans, Rosie Perkins, David McDaid, Roz Shafran, Alexandra Pitman, Sonia Johnson

**Author notes:** **Author contributions** All authors contributed to the study design, data analysis, interpretation of findings and writing up.

## Abstract

**Background:** Many mental health conditions are associated with loneliness, which is both a potential trigger and an exacerbating factor in mental health conditions. Richer evidence about how people with mental health problems experience loneliness, and about what exacerbates or alleviates it, is needed to underpin research on strategies to help with loneliness in this context.

**Methods:** Our aim was to explore experiences of loneliness, as well as what contributes to or helps address it, among a diverse sample of adults living with mental health problems in the UK. We recruited purposively via online networks and community organisations. Qualitative semi-structured interviews were conducted with 59 consenting participants by video call or telephone. Researchers with relevant lived experience were involved at all stages, including design, data collection, analysis and writing up of results.

**Findings:** Analysis led to identification of four overarching themes: 1. What the word “lonely” meant to participants, 2. Contributory factors to ongoing loneliness, 3. Connections between loneliness & mental health, 4. Ways of reducing loneliness. Central aspects of loneliness were lack of meaningful connections with others and lack of a sense of belonging to valued groups and communities. Some drivers of loneliness, such as losses and transitions, were universal, but specific links were made between living with mental health problems and being lonely. These included direct effects of psychiatric symptoms, the need to withdraw to cope with mental health problems, and impacts of stigma and poverty.

**Conclusions:** The multiplicity of contributors to loneliness that we identified, and of potential strategies for reducing it, suggest that a variety of approaches are relevant to reducing loneliness among people with mental health problems, including peer support and supported self-help, psychological and social interventions, and strategies to facilitate change at community and societal levels. The views and experiences of adults living with mental health problems are a rich source for understanding why loneliness is a frequent experience in this context and what may address it. Co-produced approaches to developing and testing interventions have potential to draw on this experiential knowledge in formulating effective approaches to loneliness.

## Introduction

Interrelationships between loneliness and mental health problems are the focus of a growing body of literature (Lim et al. 2020). Loneliness is defined as a subjective experience where individuals feel a discrepancy between social relationships that they desire to have and the social relationships that they actually have (Perlman & Peplau, 1981). Loneliness is more prevalent among people with mental health problems than in the general population (Lauder et al 2004, Badcock, 2015; Lim et al 2020).

Potentially important associations have been found between loneliness and a range of health indicators. It is a risk factor for multiple poor physical health outcomes, including early mortality, impaired cognition, hypertension, stroke, and cardiovascular disease (Petitte et al, 2015; Hawkley et al, 2010; Steptoe et al, 2013; Valtorta et al, 2016). Health service use is also higher among lonely people, especially older people (Gerst-Emerson et al, 2015; Newall et al, 2015). Regarding mental health, loneliness appears to put people at risk of onset of depression (Mann et al, 2021; Lee et al. 2021), whilst loneliness (and a closely related construct, lack of subjective social support) is associated longitudinally with recovering less well poorer recovery from mental health problems (Wang et al, 2018, Ma et al. 2021).

Whilst the evidence linking loneliness to mental ill health, and its inherently distressing nature, make loneliness a potentially promising focus for developing strategies to improve quality of life and outcomes among people living with mental health problems (Mann et al. 2017), few evidence-based and implementation-ready interventions are available (Ma et al. 2020). Strategies to reduce loneliness are more likely to be successful if rooted in an understanding of what people with mental health problems mean when they say they are lonely, how this relates to experiences of mental distress, and what they find improves or exacerbates loneliness. Much of the empirical research on loneliness and mental health has deployed measures that treat loneliness as a straightforward uni-dimensional phenomenon. This is despite philosophical, historical and experiential writing suggesting that the term captures a complex cluster of emotions and experiences (Svendsen, 2021; Alberti, 2021). Currently used measures are also not tailored to investigating loneliness in the context of mental-ill health, nor have they been developed in collaboration with people with relevant lived experience.

Qualitative research on lived experiences of loneliness among people with mental health problems has important potential to yield a richer account of the nature of such experiences and what improves or exacerbates loneliness. Such an understanding should underpin further research, including development of interventions, measures and hypotheses for quantitative investigations. The few published investigations are small in scale, and suggest a complex, intertwined relationship, with loneliness both exacerbating and arising from mental health problems. A phenomenological study of people diagnosed with “borderline personality disorder” (Sagan, 2017) found that the loneliness experienced amongst this group of participants was perceived as rooted in traumatic early experiences and strongly associated with negative feelings about self and others. Participants also described feeling disconnected from those around them and on the outside of social activities at which they were present. Lindgren and colleagues (2014) conducted a qualitative study with five individuals with mental health problems and participants described multifaceted and shifting experiences of loneliness that varied with life situation but were also very persistent. A meta-synthesis of studies on the experience of loneliness among young people with depression identified a range of factors, including depressive symptoms, non-disclosure of depression, and fear of stigma, which perpetuated cycles of loneliness and depression (Achterbergh et al, 2020). However, the qualitative literature on loneliness experiences among people living with mental health problems remains overall very limited in scope and size.

Our aim in the current study is therefore to develop an understanding of the lived experiences of loneliness among a broad range of people living with mental health problems. This was identified as a high priority evidence gap by the UKRI Loneliness and Social Isolation in Mental Health Research Network, a cross-disciplinary research network established to advance research on the relationship between loneliness and mental health (Pitman et al. 2018).

## Methods

We conducted this qualitative interview study using a co-production approach, involving collaboration between people with relevant lived experience, clinicians and university-employed researchers (some of the team had multiple relevant perspectives). Semi-structured individual interviews were used to explore the experiences of loneliness in adults with mental health problems.

Ethical approval for the study was obtained from the UCL Research Ethics Committee on 19/12/2019 (ref: 15249/001). Ten interviews were conducted prior to the March 2020 onset in the UK of the COVID-19 pandemic. When the pandemic-related restrictions began, a decision was made in conjunction with the NIHR Mental Health Policy Research Unit team (https://www.ucl.ac.uk/psychiatry/research/mental-health-policy-research-unit), to extend the study to include experiences of the pandemic among study participants. This allowed us to gather data both regarding our original research question on loneliness and regarding the impact of the pandemic on people already living with mental health problems, a topic on which evidence was urgently needed early in the pandemic. An ethics amendment was therefore submitted and approved on 04/05/2020, to conduct the interviews online and use an amended topic guide also covering experiences of COVID-19. Findings relevant to experiences of loneliness are reported in this current paper; we report the material on the pandemic elsewhere (Gillard et al. 2021, Vera et al. 2021; Shah, Hardy et al 2021).

### Research Team

A team of Lived Experience Researchers (LERs) and researchers from the UKRI Loneliness and Social Isolation and Mental Health network planned the study. Shortly after the onset of the pandemic, the team was augmented by NIHR Mental Health Policy Research Unit (MHPRU) researchers, including members of the Policy Research Unit’s Lived Experience Working Group. The MHPRU, a unit funded to deliver evidence rapidly to address policy needs, was able to contribute additional resources to extend the study to include experiences during the COVID-19 pandemic.

The research team met weekly by Zoom (Zoom Video Communications, Inc., 2020) to plan the study and discuss progress. The team included clinical academics and non-clinical researchers from a range of backgrounds (including qualitative and mixed methods research, health policy, health economics, and the arts), some of whom were members of the UKRI Cross-Disciplinary Research Network on Loneliness and Mental Health.

MB, a researcher with extensive experience of conducting qualitative research, and an occupational therapist, conducted all the telephone interviews. Thirteen LERs conducted the face-to-face and online interviews, and all had personal experiences of using mental health services and/or mental distress. Three LERs were employed in university research roles; others were members of research advisory groups and had honorary research contracts with University College London. Eleven of the LER interviewers were female and five were from minority ethnic backgrounds.

The LERs received training regarding conducting face-to-face and online interviews and obtaining written and verbal informed consent. A weekly lived experience reflective space provided LERs with emotional support and space to discuss the research process and emotional impact, peer-facilitated by four experienced LERs.

### Sampling and recruitment

Purposive sampling was used to ensure diversity regarding participants’ mental health diagnoses, use of mental health services, and demographic characteristics, including age, gender, ethnicity, and sexuality, and whether they lived in rural or urban areas. We reviewed our sample characteristics weekly during recruitment and implemented targeted strategies to ensure diversity. These included approaching community organisations with a specific remit to work with Black and Minority Ethnic communities and the use of targeted recruitment materials in an attempt to increase participation amongst these communities.

Participants were eligible to take part if they were aged 18 years or over and had a self-reported mental health problem, and lived in the UK. Three London-based community organisations (a mental health charity, a homeless charity and a community arts organisation) agreed to host the interviews prior to March 2020, although we were able to conduct only two face-to-face interviews prior to onset of the COVID-19 pandemic. Following the pandemic onset and the switch to a more extensive study in online form only, we publicised the study on the social media platform Twitter via a range of personal and organisational accounts, including with the support of the Mental Elf blogger (The Mental Elf (nationalelfservice.net). Several charities and community organisations supporting people with mental health problems also agreed to disseminate an invitation to participate to those accessing their organisations.

Potential participants contacted the research team by email. Researchers then checked eligibility, provided a participant information sheet, answered questions about the study, and booked interviews.

### Data Collection

The topic guide (see Appendix 1) was initially developed collaboratively by members of the Loneliness Network’s Co-Production Group, which includes researchers from the Network. Questions and prompts were designed to address our research questions regarding the nature of loneliness experiences and alleviating and exacerbating factors. Following the onset of the COVID-19 pandemic, questions were also added regarding experiences of living with mental health problems during this pandemic (see Appendix 2 for amended topic guide).

The interviews were conducted face-to-face (prior to the pandemic), via telephone and online via Microsoft Teams video-conferencing software. Participants had the option of joining using a freephone number so those without access to a computer could take part in an interview. The interviews took place between March and July 2020. Lived Experience Researchers conducted two face-to-face interviews, and all of the 49 online interviews, with a second researcher also present during the online interviews to manage technical aspects of data collection. Eight telephone interviews were conducted by MB before approval of the ethics amendment to conduct the interviews online. Written informed consent was obtained for face-to-face interviews. For telephone and online interviews each item on the consent form was read out loud by the interviewer and verbal consent gained for each item, which was audio-recorded. The video files of interviews were converted to audio files prior to transcription internally, or by an external transcription company where participants agreed to this). Interviews were anonymised following transcription.

### Analysis

We took a participatory approach as a large team of researchers from a range of backgrounds. Data were analysed using Template Analysis (King and Brooks, 2017) a form of thematic analysis (Braun and Clarke, 2019) that uses a codebook approach. Template Analysis involves defining and organising themes using a coding template used by those involved in analysis of the transcripts, and the template is developed and refined using an iterative process during data analysis.

Analysis of the interview transcripts was carried out by four Lived Experience Researchers (RRO, BC, PS and PN) and a network researcher (MB). A qualitative data analysis software package, NVivo (QSR International Pty Ltd., 2018) was used to manage the data. MB initially analysed three transcripts by reading and re-reading the transcripts and identifying initial themes, and three LERs also independently analysed one each of those transcripts, and discussed points of divergence with MB. Differences were compared to highlight any areas that needed closer examination in analysis of future transcripts and to ensure the coding captured complex and nuanced data relevant to the research topic. The aim was to capture richer data, to guide further coding, and not to seek a consensus on meaning, in keeping with the approach of reflexive thematic analysis (Braun and Clarke, 2019). All themes formed an initial coding template. A further set of five transcripts were then analysed individually and the further initial themes were then discussed between the five people conducting the analysis. This group reflected on the themes together to ensure important ideas in the transcript had not been overlooked, and to refine the initial themes in the coding template.

Throughout this process the core coding team met fortnightly with a wider team of Network researchers and Co-Production Group members to discuss the list of themes in the coding template as they developed, to broaden perspectives on the analysis. These meetings progressed the iterative development of the list of themes in the coding template, defining, naming and reorganizing them through discussion.

### Results

A total of 59 participants took part in the study. Most of the participants were female (n=41, 69%), aged between 25-54, and living in a city (n=43, (73%). The main ethnic groups reported were White British (n=32, (54.2%), White Other (n=8, 13.5%) and Black/Black British (n=7 (11.9%). Ten participants were interviewed during our initial (pre-pandemic) phase of recruitment (two face-to-face, 8 via telephone) and 49 participated in online interviews following the pandemic’s onset (with interviews also including questions on COVID-19 and mental health). Table one presents a summary of the demographics of participants.

**Table one:** Demographics of 59 participants

|  |  |
| --- | --- |
| **Gender** |  |
| Female | 41 (69%) |
| Male | 18 (31%) |
| **Age** |  |
| 18–24 | 2 (3.4%) |
| 25–34 | 15 (25.4%) |
| 35–44 | 15 (25.4%) |
| 45–54 | 16 (27.1%) |
| 55–69 | 5 (8.5%) |
| ≥ 70 | 4 (6.8%) |
| Info not avail | 2 (3.4%) |
| **Ethnicity** |  |
| White British | 32 (54.2%) |
| White Other | 8 (13.5%) |
| Mixed/multiple ethnic groups | 5 (8.5%) |
| Asian/Asian British | 6 (10.2%) |
| Black/Black British | 7 (11.9%) |
| Other Ethnic Group | 1 (1.7%) |
| **Sexual orientation** |  |
| LGBTQI<sup>a</sup> | 9 (15.3%) |
| Heterosexual | 43 (72.9%) |
| Prefer not to answer or information not available | 7 (11.8%) |
| **Self-reported diagnosis** |  |
| Mood disorders (depression, anxiety, PTSD) | 26 (44%) |
| Personality disorder | 7 (12%) |
| Bi-polar disorder | 8 (13.5%) |
| Schizophrenia/psychosis | 7 (12%) |
| Other (addictions, suicidal thoughts, OCD) | 6 (10%) |
| Not stated | 11 (18.5%) |
| **Current or past use MH services** |  |
| None (n =13), or on waiting list (n = 3) | 16 (27.1%) |
| Inpatient services<sup>b</sup> | 3 (5.1%) |
| NHS community mental health services<sup>c</sup> | 28 (47.4%) |
| GP or primary care counselling | 5 (8.5%) |
| Private sector psychotherapy only | 2 (3.4%) |
| Voluntary sector mental health services only | 4 (6.8%) |
| No information available | 1 (1.7%) |
| **Urban/Rural location** |  |
| City | 43 (73%) |
| town | 12 (20.3%) |
| Village | 3 (5%) |
| No Information available | 1 (1.7%) |

<sup>a</sup>LGBTQI included: gay (n = 1), lesbian (n = 1), bisexual (n = 6), pansexual (n = 1)
<sup>b</sup>Inpatient services included: acute inpatient ward (n = 2), crisis house (n = 1)
<sup>c</sup>Community mental health services included: community mental health team (n = 22), reablement team (n = 1). Therapist (n = 3), NHS peer support service (n = 2)

Our analysis identified a final set of four themes, of which themes one and four captured responses to specific questions asked in the interviews.

### Theme One: What the word “lonely” meant to participants

Participants described loneliness in a variety of ways, including reference to emotions or physical sensations experienced, anecdotes portraying loneliness, and their connection to other things and people.

#### 1.1 How Loneliness Feels Emotionally

Participants often used words synonymous with low mood including sad, depressing, miserable, despondent, tearful, and generally they conveyed that loneliness negatively affected their mental health. Other participants used terms relating broadly to feelings of separation or thwarted belongingness, e.g., longing, disconnected, “on the outside looking in”, and “missing someone or something, but not always knowing what.”

#### 1.2 Not Feeling Connected

Being alone featured in definitions, but the experience of not feeling connected was more prominent than depictions of physical solitude:

“*The last 35 years has been lonely*…*I mean I’ve got a husband, I’ve got four kids and everything else but it doesn’t stop you from feeling lonely*.*”* [P47, female, White British, 46-55, urban]

People spoke about emotional, psychological and spiritual disconnection and “*not sharing energy”* [P9, female, White other, 36-45, urban].

It was not always clear when the sense of disconnection had begun but there was an awareness that something had been lost or was missing:

“*Unable to connect* …*on an emotional level*” [P44, female, White British, 36-45, urban]

“*it’s more of a psychological state of not feeling connected to the social world*.*”* [P20, female, White British, 26-35, urban]

Participants frequently described being in the company of others yet feeling unable to connect with them emotionally. This was a chronic state for some, or coincided with periods of mental unwellness for others:

*“Remember becoming very depressed and* …*with a group of friends doing things I used to enjoy and literally not feel any connection with the people around me, especially if I was having a lot of intrusive thoughts*…*”* [P44, female, White British, 36-45, urban]

Feeling disconnected seemed often to be described more as an inner state of being than a reflection of external circumstances. Some of these statements seemed to conceptualise loneliness as something that went beyond relationships merely with people: *“Disconnect from the world, both the physical and the spiritual world as well*.*”* [P37, female, Black, 26-35, urban]

*“Not connected to anything else that’s like living or sentient*.*”* [P46, Female, White British, undisclosed, urban]

#### 1.3 Choice and Control

Some participants recognised an element of choice in electing not to communicate with people or to exist in self-imposed isolation, yet also paradoxically described how they felt lonely: “*Lonely means*… *not communicating with people and distancing yourself from society”* [P7, male, Asian British, 26-35, urban]

*“I could pick up the phone and ring someone anytime any day and I don’t…so that is sort of self-imposed isolation*.*”* [P11, male, White British, 66-75, urban]

One participant, who had experienced childhood trauma, said they avoided authentic connections, and always ‘wore a mask’ when relating to others because that kept them safe: “*There’s certain parts of you, you just don’t let people in. And that can be lonely*.” [P47, female, White British, 46-55, urban]

Choice and control, however, were complex, and beneath the conscious decisions there could be unconscious barriers or mental health factors:

*“It could be something which is self-inflicted for example, when I was depressed I felt lonely but some of it was that I felt comforted by being left alone and yet I also felt really anxious about being alone as well*.*”* [P49, male, British Asian, 46-55, urban]

*“I have less power over the thing that’s causing me to be alone or to feel alone*.” [P33, female, white British, 26-35, urban]

#### 1.4 Being or Feeling Alone

For some people, loneliness stemmed from being in a context or environment where they felt unable to connect or relate to those around them and share interests and values. For one female participant this feeling of being ‘other’ followed relocation from a large city to a more rural area with a different recreational culture:

*“It’s not actually the act of being alone, it’s*… *not having [the right] people to do things with or to share things with*.*”* [P2, female, 36-45, White British, urban]

The number of people and quality of interactions were also factors:

*“I feel lonely. because there’s no one around me apart from my mum when I’m at home*.*”* [P38, male, Black, 46-55, urban]

Loneliness could simply be defined as missing one specific person, articulated by [P31] who was living in a different city to their partner. Others described objectively being isolated: *“Some people*… *they have no choice*… *they don’t have the care and support from family and friends*.*”* [P38, male, Black, 46-55, urban]

Contextually the interviews occurred under COVID lockdown when many social and mental health communications were digital. Physical presence clearly mattered to many people. One participant described loneliness as:

“*Less connection to people and specifically decreasing face-to-face contact*.*”* [P28, female, White British, 56-65, urban]

Pets were mentioned by several interviewees. Their status as valued companions sometimes appeared on a par with humans, and their constancy was an asset that could be both cherished or missed:

“*Not having contact with other people or animals”* [P49, male, British Asian, 46-55, urban]

#### 1.5 Not feeling loved or having emotional needs met

Not feeling loved or supported underpinned many of the narratives, sometimes resulting in a deep sense of emotional pain. This applied whether participants were objectively isolated or not:

“Don’t have anyone to speak to or anyone to support you… don’t have anyone to turn to.” [P34, female, Asian, 18-25, urban]

Expressing doubts about whether health services or other people actually cared was common: “*No one helps, no one cares*.*”* [P4, female, White British, 26-35, urban]

Several interviewees recounted social rejections that had triggered self-doubts, and these ruminations undermined their mental wellbeing:

*“You think*…*this person is not interested in me. Then you start to* … *make some negative judgements about yourself* “[P10, male, Black British, 56-65, urban]

#### 1.6 Not feeling understood

Not feeling understood was one factor that increased feelings of loneliness:

*“When I’m not feeling understood*… *it feels like I’m on my own little planet*.*”* [P9, female, White other, 36-45, urban]

Sometimes the absence of understanding related specifically to mental health issues, especially where people felt they could not be open about their difficulties:

*“Because I didn’t think anyone else was experiencing the same sort of stuff as me, I wasn’t open about my mental health*…, *so I wasn’t able to connect with people in that way and felt very lonely*.*”* [P5, female, White British, 26-35, rural]

For some, loneliness related to cultural taboos about concepts of mental health. Being misunderstood by those people closest to them could result not only in feelings of loneliness but of being ostracised:

*“People within my culture*… *my family don’t understand and friends from my culture don’t get it so I feel more isolated. I’m losing friends because of this”* [P34, Female, Asian, 18-25, urban]

*“Feeling like you’re the only one*.*”* [P18, female, British Asian, 36-45, urban]

#### 1.7 Physical Sensations of Loneliness

A small number of participants described loneliness as physical sensations, such as “*tightness or stabbing in the heart”*, and *“a body ache”*, but with most sensations relating to the digestive system, namely as *“nausea”, “hunger”, “craving”* or *a “blow in the guts”*. “*Physically feel nauseous*.*”* [P22, male, White British, 26-35, urban]

*“A stabbing in my heart*… *maybe that stabbing in the heart is this thing of being rejected?”* [P6, female, White other, 66-75, urban]

There was no clear pattern to these somatic sensations.

##### Theme Summary

Overall, the data coded under Theme One showed that the experience of loneliness is complex and multi-faceted with many intertwined factors. Although these interpretations of loneliness were diverse, a unifying element was that loneliness was far from just being an unwanted lack of company, but had important psychological elements.

### Theme Two: Contributory factors to ongoing loneliness

#### 2.1 Root causes of loneliness

Participants described their perceptions of the root causes of loneliness. The internal causes of loneliness described included being an introvert or having low self-esteem and self-confidence, and therefore spending less time around people:

*“Because I have issues of self-esteem and all that kind of thing, low self-esteem, low confidence, I unintentionally do things that make me feel more lonely. For example, I don’t go to birthdays and that kind of thing. Which causes me to feel lonely*” [P41, female, Black British, 26-35, urban]

Participants also described how past experiences of being bullied negatively impacted how they viewed their social interactions. In this way, external causes of loneliness such as earlier trauma influenced thought processes and choices which became internal causes of loneliness in the present:

*“Sometimes I feel comfortable to just go up to them* [at work function] *and interrupt join in the conversation but other times I feel like ‘oh they haven’t included me therefore they don’t want me there’* …*and I think that has probably stemmed from you know being bullied at school. …*.*and then feeling like I’m being left out deliberately because that’s how the bullying worked at the time*.” [P21, female, White other, 36-45, urban]

Some participants referenced earlier traumatic events such as bereavement or domestic violence as relevant to their loneliness because these were times of great deficit in terms of having one’s emotional needs met. There were painful memories of the absence of support, love and care within the family home which may have affected childhood emotional development:

*“I’ve kind of lived in a household that didn’t really know how to show love. So there was a lot of violence…. I hate being on my own…. Stayed in relationships when I probably shouldn’t have when I was younger. That was violent*.*”* [P9, female, White Other, 36-45, urban]

#### 2.2 Current factors that maintain loneliness

Participants described a range of current factors that influence their loneliness. These included separation and loss, and current relationships and the impacts of these on their mental health conditions. A sense of not belonging in communities, including neighbourhoods, workplaces and other group settings, was also experienced as an important driver of loneliness. People not feeling they belonged often seemed to be characterised by a complex interplay of overlapping characteristics, including having a mental health condition and also disability, race or social class.

##### 2.2.1 Loss or separation from meaningful relationships

Forms of loss that resulted in loneliness included romantic, platonic, physical and emotional loss, encapsulated in participants’ accounts of loss of friendships and of love, and loss through bereavement. One participant felt that their independence and well-being, especially their mental health, required them to stay physically away from family and friends, but that this nonetheless resulted in a sense of loneliness:

“*The one thing that I need for my sanity is my own space*…*although it’s better for me to live here* [in own home far from family and long-term friends] *because I have that, I do feel lonely*.’’ [P2, female, 36-45, White British, urban]

For some, grief was a factor in feelings of loneliness and how it currently limited them in doing other activities which might have reduced their feelings of loneliness:

“*I have faced grieving for someone that died close to me. I despaired and just be at home alone and just not able to function…*’’ [P48, female, 46-55, White other, urban].

##### 2.2.2 Compounding intersectional factors and external barriers

For some with a mental, physical or age-related disability, associated barriers to using transport and getting involved in activities contributed to current feelings of loneliness. As one participant noted:

“*When I developed a physical disability, I found there was very little in my community as a person in my thirties that I was eligible to do*.*”* [P44, female, White British, 36-45, urban]

For this woman the problems were structural in that access to activities might be restricted by age range, or preconceptions about the type of activities that it was perceived that any certain demographic would be interested in or capable of.

Even within a group that you “belong” to there could be differences in interests, attitudes, speech style, access and ability and social norms. Other compounding intersectional factors cited by participants included sexuality, discrimination, race relations and the complexities of age differences in interactions with others.

##### 2.2.3 Time Alone – impacts of solitude

Some participants described the negative impact spending time physically alone had on mental health and how it related to feelings of loneliness:

“*I get very disconnected from what’s going on and I struggle to keep my thoughts logical… bordering on psychotic but not quite psychotic. Quite dissociated*.*”* [P46, female, White British, age not known, urban]

“*It will affect my mood, it will make me feel low. It will affect my loneliness greatly if there are several days I have not been out somewhere or seen someone or had face-to-face time with people*…*”* [P23, male, Asian British, 36-45, urban]

Some participants described how time alone led to mood changes or harmful behaviours: “*Every time… I’ve behaved in self-destructive ways, it’s been when I’ve been on my own. My thoughts can very quickly spiral out of control in unhelpful ways*” [P1, female, White British, 36-45, urban]

“*It’s hard for me to be by myself in a healthy way*… *I get cranky. And it kind of impacts on itself, and I get crankier. And I feel less able to say how I feel*… *disconnected from my feelings*” [P40, male, White British, 46-55, rural]

The negative effects of time alone included a loss of social skills and the difficulties this created in interacting with others:

“*You forget how to socialise. You become kind of quite selfish actually, you don’t think about other people’s feelings*.” [P42, female, Asian, 36-45, urban]

However, some participants described a need to withdraw socially and ensure periods of solitude, even though this might exacerbate loneliness:

“*I need time on my own just to recover and relax and rejuvenate but I don’t think it’s good to be on my own for long periods of time*” [P8, male, White British, 36-45, urban]

##### 2.2.4 Sense of belonging

Sense of belonging (or not belonging) to a community or a group was interpreted in various ways, with a number of participants describing ways in which this affected their loneliness: “*No, I don’t feel part of this community, not at all*…*I feel lonely where I live yes*. “[P51, female, White other, 46-55, urban]

Additionally, social class played a role in some individuals’ sense of not belonging, especially as evidenced in their experiences of being in the workplace with people from a different background to them:

*“I’m from a working-class background, I’m very around middle-class professionals at work who’ve had-who speak very differently to me. …I have to constantly think about how I’m coming across, how I’m speaking… makes me feel a bit disconnected…I rarely join them for lunch and things … I find it a bit like I’m an outsider*” [P37, female, Black African, 26-35, urban]

One participant reflected on her constant efforts to find a sense of belonging:

*“I feel like I struggle with my identity sometimes, um, like who am I and stuff like that. Yeah, I’m always like, trying to find like, my tribe, and like-minded people and stuff like that*.” [P29, female, Black, 26-35, urban]

Similarly, one white British male also described a sense of not belonging in his community because of having a mental health condition:

*“Sometimes the feelings of, well I do belong to that community but do I really? Am I a bona fide member of it? Feel like I am a hanger-on in the community*.” [P11, male, White British, 66-75, urban]

Conversely, some people described positive neighbourly encounters and informal interactions within a local community that increased some participants’ sense of belonging and lessened their feelings of loneliness.

One participant had lived in a multiple occupancy house and had found that despite their struggles with their mental health, being in proximity to others had helped build friendships that increased their sense of belonging:

*“I had people that were close… in proximity quite a lot, and we got to become really good friends, and I felt safe in that sort of friendship and then was able to open up, even when I wasn’t feeling great”* [P5, female, White British, 26-35, rural]

Furthermore, connections through activities in the community helped some participants overcome feelings of loneliness and made them feel as though they belonged:

“T*he gym was really great, it was quite near me you know, and I’d never been to the gym before but I really got into that, and I was just getting to know people’’* [P6, female, White other, 66-75, urban]

Similarly, another participant described participating in group activities as part of their academic studies as countering loneliness through a feeling of belonging:

*“The last time I remember not feeling lonely was when I had my lectures and going into my lectures because it was more like group workshops on my course so that was good for me*.*’’* [P34, female, Asian, 18-25, urban]

Thus, a feeling of belonging could be due to activities or situations that were more enforced than freely chosen, such as sharing housing, or being tasked to do a group activity in an academic setting. However, some participants actively sought opportunities that would enhance a sense of community through engaging in activities they felt passionate about:

*‘‘I managed to find the live role-playing community*… *I have organised events to help kind of increase the hobby outreach and activities within the hobby*.*’’* [P19, female, White British, 36-45, urban]

For this participant, finding her kind of people online led to her then taking the initiative and organising her own online events and thus wrapping this community around her.

##### Theme summary

Overall, the data under theme two captured a range of root causes of loneliness, in addition to current factors that cause and contribute to loneliness. The data also demonstrated cross-cutting relationships between a sense of not belonging to a group or community, one’s mental health and feelings of loneliness. Participants described how the sense of belonging through community or identity impacted positively on their loneliness and mental health. Others described how not feeling they belonged had a negative impact on their mental health and feelings of loneliness.

### Theme Three: Connections between loneliness & mental health

Participants described close connections between their mental health and feeling lonely. These included both loneliness leading to deterioration in mental health, and conversely feeling lonely as a result of living with a mental health problem. Some participants were aware of this as an apparently cyclical relationship and for others it was a single pathway in one direction.

#### 3.1 Loneliness exacerbating mental health problems

Social isolation and loneliness may mean that people are less able to access support from people in their immediate environment. Having someone around could serve as psychological reassurance, or as a sounding-board and reality check for people which could, for example, prevent a spiral into psychosis. Being alone with one’s thoughts can be detrimental. Some participants described how ruminating on feeling lonely impacted negatively on their mental health:

*“I feel lonely then I start thinking that I don’t really have people to talk to or anyone to support me then I start feeling depressed, I feel really low, a lot of depression*.*”* [P34, female, Asian, 18-25, urban]

For this participant rumination led to a progression of negative thoughts, undermining their mood and exacerbating the problems.

#### 3.2 Mental ill-health leading to loneliness

Participants also described how the symptoms of their mental health problem contributed to loneliness. This could involve isolating themself at home, and or changes noticed by friends, leading to loss of social contact and loneliness over time:

*“My depression involved me sitting in the dark at night, with lights off [laughs]. I couldn’t sleep, eat. I used to be popular. People stopped coming to see me because I’d changed. My personal hygiene was not the best [laughs]. Yeah so I became lonely then in the end*” [P10, male, Black British, 56-65, urban]

In the end this participant felt there was no choice but to be on their own, as they no longer had contact with friends, and anxiety reduced their ability to reach out to people:

*“After* [developing depression] *I just got comfortable being on my own. I wouldn’t say comfortable. There was no other alternative, was there? If people are not coming to see you, you haven’t got…you get anxious going to see other people, there is no alternative but to be on your own*.*”* [P10, male, Black British, 56-65, urban]

Participants also described the impact of stopping activities, as a result of depression, as contributing to their loneliness:

*“If I withdraw from doing things due to you know, feeling depressed and not enjoying them then I become more lonely and it’s a bit of a downward spiral*” [P44, female, White British, 36-45, urban]

Some participants identified difficulties they associated with their specific mental health problems, for example, challenges in making and maintaining relationships, as resulting in loneliness:

*“Because of my condition* [diagnosed with “borderline personality disorder”], *I worry about having relationships with people and that keeps me isolated and keeps me lonely”* [P1, female, White British, 36-45, urban].

The impact of hearing negative voices was also identified as limiting interactions:

*“Getting out the flat is hard because of mental health and then having conversations with friends, like the voices would tell me stuff like ‘they hate you’ ‘don’t talk to them’ so that side. And then sort of like lacking motivation sometimes to sort of hold conversation*.*”* [P20, female, White British, 26-35, urban]

Some participants reported that experiencing anxiety whilst in a social situation resulted in feeling disconnected from others and lonely despite being in company:

*“So anxiety can make me feel kind of lonely even if I’m in a party. I can be in a party with all my friends, all just want me to be happy and free and relaxed and dance, enjoy the music. But I can’t come out of my shell. I’m constantly preoccupied with my anxiety and this sense of…not enjoying myself. This sense of fear. And there is a resolve of not enjoying myself and having to behave like I am enjoying myself. Then I do kind of feel very lonely because it looks to me like everyone else is enjoying themselves whereas I’m not*.” [P8, male, White British, 36-45, urban]

Some participants described that living with a mental health problem such as anxiety and resultant avoidance of social situations, eroded social contacts over time:

*“I’ve always been suffering with anxiety, so… being alone was easier*… *but I am quite a social person…as I’ve got older* …*the anxiety has got worse”* [P26, male, White British, 36-45, urban]

Some participants found managing day-to-day life tasks, such as handling household chores, in addition to coping with their mental health problems challenging, resulting in less time available to address their loneliness:

*“Always playing catch up to get everything sorted in my life, but it becomes fairly low down in my list to sort of join groups or develop hobbies. I think …I have to find more ways to either quickly execute the things that need to be done to run a life and a flat or be very strict about just cutting things that don’t seem to matter so much and so concentrate more on*…, *finding ways to sort of overcome loneliness”*. [P28, female, White British, 56-65, urban]

#### 3.3 Cyclical relationship

Participants described the relationship between feeling lonely and changes in their mental health as a cycle, perpetuated by difficulty in connecting with people:*”It’s a bit of a vicious cycle because I think feeling lonely- and because we’re social animals, feeling lonely will make it worse, you need to be able to talk to people*… *So it’s a bit of a cycle, so your mental health means you can’t connect to people, then not being able to connect makes your mental health worse and then you’re just cycling around*.*”* [P46, female, white British, undisclosed, urban]

#### 3.4 Stigma

A key external factor reported by several participants was the negative impact of stigma related to their mental health problems on relationships:

“*People walk away. You mention the word hospital, people walk. You mention the word psychiatry and people walk away*.” [P50, male, White British, 56-65, urban]

“*I think because of what I have, and the stigma associated with personality disorders* … *in the press*, …*some of the family members, some work colleagues, I think if I’m feeling lonely then that sort of, all that sort of stigma, I think it just all compounds more”* [P30, female, White British, 36-45, urban]

##### Theme Summary

Overall, the data under theme three showed that loneliness and participants’ mental health, and indeed the stigma associated with their mental illness, were intertwined and influenced each other in a variety of ways.

### Theme Four: Ways of reducing loneliness

Participants shared what had helped them to feel less lonely and what advice they would offer to others. Two broad sub-themes emerged - external and internal approaches. These were categorized based on participants’ emphasis and experiences, but were often intertwined rather than being two clearly distinct categories. Strategies and ideas classed as “external ways to reduce loneliness” are those where participants emphasised how their experience depended on specific needs being met by people or environments external to themselves; those classed as “internal ways to reduce loneliness” highlight internal changes in thought patterns or other psychological shifts in participants’ ways of being and relating to the world around them.

#### 4.1 External ways to reduce loneliness

Many participants described how social contact was key to helping them feel less lonely. Some people had a preference for the type of contact, whereas for others, any contact with people was important:

“Try and communicate with people. Even if it’s going to Kew Gardens by yourself, you’re going to meet people when you sit down at the café and they’re going to say to you “isn’t the weather lovely?” [P38, male, Black, 46-55, urban]

The quality of connection, and being able to talk about how you were feeling was essential for many:

*“My advice would be to talk to*…*someone that you trust*…*if you say it out loud*…*it does lift your feelings a bit because you’ve connected with someone, you’ve shared, you’ve*…*offloaded to them. And someone*…*being empathetic towards you definitely helps*.*”* [P32, female, Asian, 26-35, urban]

Accessing mental health support was cited by some as a key first stage in decreasing loneliness, by helping to improve mood and anxiety, and being able to reflect on oneself and develop better coping skills:

*“Before I*… *started medical treatment for my depression and anxiety I felt that being off feeling. Even if I was around people I felt separated, that there was a kind of barrier that was stopping me from being able to communicate*.*”* [P33, female, white British, 26-35, urban]

Taking part in activities and shared interests was mentioned by many participants, with the idea of “*structured socialisation”* being a route into developing new friendships or a group identity, and technology being an enabler for some. Often, the shared interests gave purpose and fostered connection:

*“Structured socialisation, it provides that structure and*… *gives you a purpose so that you feel compelled to keep going and make friends and stuff and be sociable with other people*.*”* [P5, female, White British, 26-35, rural]

*“Technology is not*…*a bad starting point to get you back out in the real world*…*like “Meetup”, you know the app for your local area?”* [P14, female, white British, 46-55, rural]

However, finding new groups or using technology was a negative experience for some people:

*“I’ve been in groups and things and I have met up with friends and things but I just, it doesn’t really work*.*”* [P47, female, White British, 46-55, urban]

*“Some of them [Meetups] are closed groups so you can’t sort of see what they’re doing, and I sort of think if you’re closed I can’t be arsed. Also, I don’t really like doing things over the internet, which is a bit of a barrier to that. Like, I don’t like putting my photo up or chatting*.” [P2, female, White British, 45-56, urban]

Some participants reflected that finding new activities and purpose required concerted effort: *“I just have this thing that, when you walk around a town*…*there’s always posters and notices up of things happening…*., *even in your local Tesco’s*…*you’ll have what’s happening in your local community*…*libraries have loads of things happening now*.*”* [P2, female, White British, 45-56, urban]

*“When I was [living] alone in a flat*…*I was much more lonely because I wasn’t tagged into those things which helped me to be less lonely*…*But we have to find these things and, if they make sense, they give us community*.*”* [P13, male, White British, 66-75, urban]

For others, just being able to get outside alone was enough in alleviating feelings of loneliness, sometimes resulting in unplanned encounters with people that reduced loneliness, as was the development of hobbies that could be enjoyed alone:

*“I made a point of trying to go out each day. Just for a walk around the village, and I’d go to the park. To the swings but it got me talking to other Mums and Dads*.*”* [P12, female, White British, 46-55, rural]

*“Finding something*…*like a hobby that connects you with something, or something you enjoy*,.. *to give a bit of routine or something else to focus on rather than*…*moping*.*”* [P32, female, Asian, 26-35, urban]

Religious worship and having a pet were also mentioned by a few people:

*“Get a dog, if it’s possible because you know, it’s much easier to interact with people in that case*.*”* [P44, female, White British, 36-45, urban]

Participants also described how volunteering gave them a sense of purpose and connection: *“The radio [volunteer job] has been life changing*…*For me that connection*…*And people listening, realising that they are connecting*…*I feel like I’m doing something good*.*”* [P9, female, white British, 36-45, urban]

#### 4.2 Internal ways to reduce loneliness

Internal strategies leading to better self-awareness, sometimes followed by taking steps to reducing anxiety and improving mood, were mentioned by many participants:

*“I’m more recognising it now in the last few years*…*that is what loneliness looks and feels like and therefore, because you can recognise it, you suddenly think oh I haven’t seen anybody for a while, I need to*…*make more of an effort now”* [P14, female, white British, 46-55, rural]

Participants reflected on relationships and learning to recognise what worked well for them and what made loneliness worse, thus improving the quality and connection in relationships:

*“I’m trying to focus on the things that I can control. With loneliness, what I can control is kind of maybe who I’m around and who I feel most comfortable with, so maybe not hanging on to these friendships that make me feel even more lonely, even more isolated*.*”* [P37, female, black, 26-35, urban]

Some people stated that acceptance of having time by yourself, and valuing carrying out activities like writing in a journal or art was helpful, in expressing yourself and developing better self-knowledge of yourself and your interests:

*“To not beat yourself up, it’s not because there’s anything wrong with you, and that spending time on your own can be good ‘cause you can do things that you can then share with people”* [P2, female, White British, 45-56, urban]

*“The hobbies that I have are doing art*… *I do a lot of writing, that helps me express my emotions, a lot of poetry, a lot of story writing, a lot of arts and crafting*…*that helps with my creativity*…*it keeps me mentally healthy*.*”* [P43, female, Asian British, 18-25, urban]

Some participants felt less lonely when connected to others of the same ethnicity:

*“The ones that make me feel less lonely, …for a long time now, are my black friends*.. *I feel more connected… I can express myself more freely without having to talk about certain things or having to express why a certain thing was racist*…*or offensive*…*they just get it*… *so ethnicity is*… *playing a massive role in terms of feelings of loneliness*.*”* [P37, female, 26-35, Black African, urban]

Participants’ thoughts and feelings about isolation could also change as a result of psychological therapies, potentially resulting in taking steps to increase connection with others: *“Through therapy and facing what I’ve been doing*…*in terms of anxiety or depression, I understand that*…*being sociable, going out and meeting people is healthy, and the longer I stay isolated…. the harder it gets”* [P26, male, White British, 36-45, urban]

##### Theme Summary

Overall, the data under Theme Four demonstrated that different forms of social contact were seen as important for many participants in reducing loneliness, with quality of relationships and a sense of “belonging” being attributes of social contact that were particularly pertinent. However, mental health challenges could act as a barrier to improving social contact and so addressing this was seen as a key step for some participants. Internal reflections and time alone also led to better self-awareness and recognition of what was valued most in relationships. For some, this led to better quality relationships and a reduction in loneliness.

## Discussion

### Main findings

Our findings conveyed how differently loneliness can be experienced by different people, as expressed in terms of emotions, or even physical sensations. Loneliness appeared to comprise psychological elements, related to people’s thoughts and feelings about themselves in relation to other people and to wider communities, and social elements, related to the impacts of everyday interactions and contacts. Participants described a range of contributing factors to the origins of their loneliness, and the ways that their loneliness was maintained. They also perceived clear links between their loneliness and their mental health, and vice versa, and sometimes a feedback loop between the two.

Our participants’ accounts, as encapsulated in over-arching Theme 1, confirm that feelings of loneliness do not simply relate to dissatisfaction with the amount of time spent with others, but, perhaps more centrally, to not feeling connected in meaningful ways to others, and to not experiencing a sense of belonging. Loneliness has long been characterised as a physically and psychologically harmful manifestation of fundamental needs for social connection not being met (Cacioppo and Cacioppo, 2012). Our findings regarding the importance of sense of belonging can be connected to investigations of loneliness from a social identity perspective, which find that having multiple valued group memberships is associated with less loneliness and greater well-being (Haslam et al. 2018). The ways in which participants described both the nature of their loneliness and its origins were diverse, congruent with quantitative findings that suggest a complex causal web underlying loneliness (Lim et al. 2020). Contributing factors identified included external factors such as losses and transitions or excessive time alone, but also aspects of personality such as introversion or lack of self-confidence, as well as the long-range impacts on ability to form relationships of trauma and adverse early experiences (Theme 2).

Some of the emerging themes illuminate aspects of loneliness relevant across populations, but a central aim of our work was to better understand loneliness among people living with mental health conditions. We found that loneliness and mental health appear intertwined in several ways (Themes 3). Participants described how feelings of not being connected to others could arise directly from a range of mental health conditions by pathways including feeling negative about self and others; withdrawing when depressed; and feeling unable to connect with others even when in company because of preoccupying social anxiety and voices with negative content impeding trust and ability to socialise. This is in keeping with findings of greater loneliness associated with a range of mental health conditions (Erzen et al, 2018; Chau et al, 2019; Loades et al, 2020; Wang et al, 2018). Some also described how loneliness could lead directly to onset or exacerbation of depression, congruent with longitudinal studies establishing loneliness as an independent risk factor for depression (Lee et al. 2020, Mann et al. 2021), and with findings of a bidirectional relationship between loneliness and depression (Nuyen et al. 2020).

Going beyond direct links from mental health symptoms to loneliness, the actions people took to cope with their mental health problems sometimes also placed them at greater risk of loneliness, such as when they withdrew from the stresses of social contacts and activities in order to recover and to ensure they had time and energy to cope with pressing practical needs. The circular relationship between loneliness and mental ill health identified in Theme 3 evoked the paradox identified by Achterbergh et al. 2020 in their meta-synthesis of qualitative studies on loneliness and depression among young people: social withdrawal to cope with mental ill health can result in loneliness that further exacerbates mental health problems.

Significant contributors to loneliness among people with mental health problems in our sample were stigma and self-stigma associated with mental health problems, especially as impediments to sense of belonging. This is in keeping with previous findings of an association between self-stigma and social withdrawal among people with severe mental health problems (Abiri et al. 2016), and of high levels of persisting mental health stigma despite a longstanding UK public anti-stigma campaign, focused primarily on common mental health problems (Henderson and Gronholm 2018). Experiences of stigma and social exclusion related to mental health intersected for many participants with social exclusion associated with being part of other disadvantaged and marginalised groups at increased risk of mental health problems, including racial and sexual minorities and people with disabilities, and with impacts of social deprivation. As Lever-Taylor et al. (2021) argue in their qualitative study of perinatal women, an intersectional focus is helpful in understanding social drivers of loneliness. Participants’ accounts of the many impediments to a sense of belonging, including stigma, reinforce a need to take a societal and community as well as individual approach to understanding loneliness. Thus, loneliness appears to result not only from individuals’ inability to connect, but also from failures of communities to welcome and integrate people living with mental health problems and from the practical barriers to connecting with others that result from poverty (Macdonald et al. 2018).

Many of the themes and sub-themes so far discussed cohere with previous literature, for example illuminating potential mechanisms underpinning epidemiological findings. To our knowledge, our exploration of the strategies and types of help people employ to combat loneliness is more novel than in the academic literature (Theme 4). We found that many participants were aware of their loneliness and its impacts, and of a variety of strategies that could help. we reflect further on implications for interventions below.

### Strengths and limitations

Our study represents a substantial contribution to the limited qualitative literature on loneliness among people with mental health problems, as described in the introduction. We recruited a diverse sample, encompassing a range of backgrounds, types of mental health problem, service use histories and locations. Although the digital platform used for most interviews will have excluded a substantial section of the population using mental health services, we did conduct some interviews by telephone to accommodate the digitally excluded. Lived experience was embedded in the study at each stage, with people with relevant personal experiences designing interview guides, conducting interviews, analysing data and writing up findings.

Limitations include that we conducted a broad-brush analysis of a large sample of interviews, and searched for commonalities across a group that was very diverse in characteristics and experiences rather than focusing in more depth on more defined groups. The majority of interviews were conducted during the COVID-19 pandemic, and we have reported baseline (Gillard et al., 2021) and follow-up (Shah, Hardy et al., 2021) findings elsewhere, but the pandemic context may have influenced accounts of loneliness.

### Research and clinical implications

The rich and complex accounts given by participants of the nature of their experiences of loneliness, and the great variety of pathways into and out of it, indicate considerable further scope for research to understand associations between loneliness and mental health. In qualitative research, many of our themes warrant more in-depth exploration, including focusing on particular groups at high risk of loneliness, or who are lonely. In quantitative research, it would be valuable to investigate further the longitudinal relationships between loneliness and mental health problems, and the extent to which contributory factors identified in our study are also reflected in epidemiological analyses. Much research on loneliness has employed single-item or brief measures not tailored to people with mental health conditions: we reflect that such measures are unlikely to capture the diversity of experiences of loneliness and links to mental health.

As yet there are very few clearly evidence-based strategies for helping with loneliness among people living with mental health problems (Ma et al. 2020). Our findings show people with mental health problems recognising and taking steps to address their loneliness. This supports the greater deployment of co-production approaches, incorporating the forms of help that people with relevant personal experience see as potentially effective. With few exceptions (Lloyd-Evans et al. 2020), interventions tested thus far have tended not to be co-produced. The diversity of pathways into and out of loneliness that people described, and of suggested strategies, indicate that a variety of approaches are potentially helpful depending on the nature and context of loneliness. These would include self-help and psychoeducation about day-to-day strategies that people have found helpful, psychological interventions focused on thoughts and feelings about others, and social approaches to help people develop meaningful connections and a sense of belonging (Mann et al. 2017).

The range of mental health-related factors triggering and perpetuating loneliness suggest benefits to developing or adapting specific strategies for this population rather than deploying strategies developed for the general population. Approaches to loneliness in a mental health context based on peer support seem rarely to have been reported, but have clear potential benefits such as in overcoming obstacles due to stigma and self-stigma, fostering a sense of belonging and supporting people with self-help strategies to reduce loneliness. Finally, our perspective in this study was on improving the support offered in mental healthcare settings, but our findings illustrated the community-level, intersectional and socio-economic drivers of loneliness in multiple ways. We suggest that addressing such drivers will have a central influence on whether levels of loneliness can be reduced among people living with mental health problems.

## Data Availability

The data are not available for now.

## Appendix 1

1. **First of all, could you tell us about who you have contact with, both face-to-face and via technology?** Interviewer Prompts:
  - For example, are there people you see each day, every week or on a regular basis?
  - Do you belong to any groups or clubs? Do you work?
  - Could you tell me about your use of technology and social media (Facebook, Instagram, Twitter, WhatsApp, Skype, apps, online forums, assistive tech etc)? **Experience of loneliness**
2. **Can you tell me what the word lonely means to you? How would you define it?** Interviewer Prompts:
  - How does it make you feel? Physically, psychologically, emotionally e.g. social anxiety.
3. Has there been a time or times in your life when you have been lonely?
4. Would you say you feel lonely in your life in general at the moment? What is/was that like? Interviewer Prompts:
  - How does loneliness impact on you day to day?
  - Are there things that you think have triggered or underlie your feeling of loneliness? E.g. age, culture, personality characteristics, difficulty fitting in, stopping work, moving location, loss of a partner or family.
  - Are there situations or times when you feel more lonely than others? Prompt: e.g. seasonal, not working. **Social contact and loneliness**
5. **Do you feel lonely when you are in the company of others?** Are there any kinds of social contact that make you feel more or less lonely? Interviewer Prompts:
  - E.g. groups, family, special friend, in person rather than technology.
  - When you meet people, do you feel that you can talk to them easily?
6. **What is the difference between meeting people, making friends and maintaining friendships?**
7. **Do you think that spending time on your own can be helpful/therapeutic?**
8. **Does spending too much time on your own become unhealthy?**
  - [If yes] - How much is too much? Where is the line? **Loneliness and mental health**
9. **Do you think feeling lonely is connected to your mental health?**
  - [if yes]: In what ways? Interviewer Prompts:
  - Does feeling lonely make your mental health worse? In what ways?
  - Do you think your mental health problems contribute to your loneliness? [Prompts: e.g. side effects of medication, the age you developed mental health problems.]
  - Do you feel that loneliness is different/distinct from your mental health problems?
  - Thinking back to when your mental health problems first started, do you think that feeling lonely came before or after?
  - [If loneliness came first]: Has your experience of loneliness changed since having mental health problems?
10. **Do you feel you belong in the community around you and with your family and friends?**
  - [If not]: Can you tell me more about this feeling of not belonging? In what ways, if any, does that relate to your feeling of loneliness?
11. **Have you ever discussed feeling lonely with family, friends or any healthcare professional?** If so, what happened next, did it help? Final questions:
12. **Are there ways in which you have tried to reduce your loneliness? What has and has not worked?** Interviewer Prompts:
  - Do you have coping strategies that you use?
  - Have you had support with this? What kind of support? Has this helped?
13. **What would not being lonely look like for you?** Interviewer Prompts:
  - How do you imagine your life would be different?

## Appendix 2 Revised Topic Guide

Title of Study: Exploring the lived experiences of loneliness and isolation with people with mental health problems during the COVID-19 pandemic in the UK.

The first five questions focused on the experiences of people during the COVID-19 pandemic and are excluded from the topic guide presented here, as the findings are not reported in this paper.

**Experience of loneliness**

6. **Can you tell me what the word lonely means to you?** How would you define it? Prompts: How does it make you feel? Physically, psychologically, emotionally e.g. social anxiety.
**7.** **Has there been a time or times in your life when you have been lonely?** Would you say you feel lonely in your life in general at the moment? What is/was that like? Prompts: Are there things that you think have triggered or underlie your feeling of loneliness? E.g. age, culture, personality style,, difficulty fitting in, stopping work, moving location, loss of a partner or family. Are there situations or times when you feel more lonely than others? Prompt: e.g. seasonal, not working.

Social contact and loneliness

8. **Do you feel lonely when you are in the company of others?** Are there any kinds of social contact that make you feel more or less lonely? Prompts: E.g. groups, family, special friend, in person rather than online or using technology. When you meet people in person, do you feel that you can talk to them easily?
**9.** **Do you think that spending time on your own can be helpful/therapeutic?**
**10.** **Does spending too much time on your own negatively affects your wellbeing?**
  - [If yes] - How much is too much? Where is the line? Loneliness and mental health
11. Do you think feeling lonely is connected to your mental health?
  - [if yes]: In what ways? Prompts: Does feeling lonely make your mental health worse? In what ways? Do you think your mental health problems or your treatment contribute to your loneliness? [Prompts: e.g. side effects of medication, the age you developed mental health problems.]
  - Thinking back to when your mental health problems first started, do you think that feeling lonely came before or after?
  - [If loneliness came first]: Has your experience of loneliness changed since having mental health problems?
12. **Do you feel you belong in the community around you?** Prompts: explore different communities: neighbourhood, family and friendship groups, communities of interest [If not]: Can you tell me more about this feeling of not belonging? In what ways, if any, does that relate to your feeling of loneliness? **Final questions**
**13.** **Are there ways in which you have tried to reduce your loneliness? What has and has not worked?** Prompts: Do you have ways of coping? Have you had any support? What kind of support? Has this helped?
**14.** **What would not being lonely look like for you?** **Prompts: How do you imagine your life would be different?**
**15.** **What tips or advice would you give someone who was struggling with loneliness?**

